# Silent Hypoxia in Coronavirus disease-2019: Is it more dangerous?-A retrospective cohort study

**DOI:** 10.1101/2021.08.26.21262668

**Authors:** Prashant Sirohiya, Arunmozhimaran Elavarasi, Hari Krishna Raju Sagiraju, Madhusmita Baruah, Nishkarsh Gupta, Rohit Kumar Garg, Saurav Sekhar Paul, Brajesh Kumar Ratre, Ram Singh, Balbir Kumar, Saurabh Vig, Anuja Pandit, Abhishek Kumar, Rakesh Garg, Ved Prakash Meena, Saurabh Mittal, Saurabh Pahuja, Nupur Das, Tanima Dwivedi, Ritu Gupta, Sunil Kumar, Manisha Pandey, Abhinav Mishra, Karanvir Singh Matharoo, Anant Mohan, Randeep Guleria, Sushma Bhatnagar

**Affiliations:** Department of Onco-anesthesia and Palliative Medicine, All India Institute of Medical Sciences, New Delhi; Department of Neurology, All India Institute of Medical Sciences, New Delhi; Department of Preventive Oncology, All India Institute of Medical Sciences, New Delhi; Department of Medicine, All India Institute of Medical Sciences, New Delhi; Department of Pulmonary medicine, critical care and Sleep disorders, All India Institute of Medical Sciences, New Delhi; Department of Laboratory Oncology, All India Institute of Medical Sciences, New Delhi; Department of Microbiology, All India Institute of Medical Sciences, New Delhi; Department of Surgical Oncology, All India Institute of Medical Sciences, New Delhi; Department of Palliative Medicine, All India Institute of Medical Sciences, New Delhi

**Author notes:** Equal contributorship (Prashant Sirohiya, Arunmozhimaran Elavarasi). **Corresponding Author** Prof Sushma Bhatnagar, Head, Department of Onco-anesthesia and Palliative medicine, All India Institute of Medical Sciences, New Delhi. **Contributorship statement** Prashant Sirohiya was involved in the conceptualization of the study, study design, patient care, and in writing the first draft of the manuscript Arunmozhimaran Elavarasi was involved in the conceptualization of the study, study design, patient care, data collection, statistical analysis, and in writing the first draft of the manuscript and agrees to be the guarantor of the manuscript taking responsibility for the integrity of the work as a whole, from inception to published article Hari Krishna Raju Sagiraju was involved in the conceptualization of the study, study design, patient care, data collection, statistical analysis, and critique of the manuscript and agrees to be the guarantor of the manuscript taking responsibility for the integrity of the work as a whole, from inception to published article Madhusmita Baruah was involved in the conceptualization of the study, study design, patient care, and in writing the first draft of the manuscript Rohit Kumar Garg and Saurav Sekhar Paul was involved in the conceptualization of the study, study design, patient care, and in the critique of the manuscript Nishkarsh Gupta, Brajesh Kumar Ratre, Ram Singh, Balbir Kumar, Saurabh Vig, Anuja Pandit, Abhishek Kumar Rakesh Garg, Ved Prakash Meena, Saurabh Mittal, Saurabh Pahuja, Anant Mohan, and Sunil Kumar were involved in study design, patient care, and in the critique of the manuscript Tanima Dwivedi, Nupur Das and Ritu Gupta were involved in study conceptualization, in analyzing the laboratory parameters, and in the critique of the manuscript Manisha Pandey, Abhinav Mishra, and Karanvir Singh Matharoo were involved in the patient care, and data collection Randeep Guleria was involved in the conceptualization of the study and final critique and review of the manuscript Sushma Bhatnagar was involved in the conceptualization of the study, convening of the working group, study design, patient care, and in final critique and review of the manuscript.

**Keywords:** Silent Hypoxia, Happy Hypoxia, Asymptomatic Hypoxia, Hypoxemia, COVID-19, Hypoxia, Coronavirus disease, SARS-CoV-2, Case fatality rate

## Abstract

**Background:** Hypoxia in patients with COVID-19 is one of the strongest predictors of mortality. Silent hypoxia is characterized by the presence of hypoxia without dyspnea.. Silent hypoxia has been shown to affect the outcomes in previous studies.

**Research Question:** Are the outcomes in patients presenting with silent hypoxia different from those presenting with dyspneic hypoxia?

**Study design and Methods:** This was a retrospective study of a cohort of patients with SARS-CoV-2 infection who were hypoxic at presentation. Clinical, laboratory, and treatment parameters in patients with silent hypoxia and dyspneic hypoxia were compared. Multivariate logistic regression models were fitted to identify the factors predicting mortality.

**Results:** Among 2080 patients with COVID-19 admitted to our hospital, 811 patients were hypoxic with SpO_2_<94% at the time of presentation. 174 (21.45%) did not have dyspnea since the onset of COVID-19 symptoms. 5.2% of patients were completely asymptomatic for COVID-19 and were found to be hypoxic only on pulse oximetry. The case fatality rate in patients with silent hypoxia was 45.4% as compared to 40.03% in dyspneic hypoxic patients (*P=0*.*202*). The odds ratio of death was 1.1 (95% CI 0.41-2.97) in the patients with silent hypoxia after adjusting for baseline characteristics, laboratory parameters, treatment, and in-hospital complications, which did not reach statistical significance (*P*=0.851).

**Interpretation:** Silent hypoxia may be the only presenting feature of COVID-19. Since the case fatality rate is comparable between silent and dyspneic hypoxia, it should be recognized early and treated as aggressively. Since home isolation is recommended in patients with COVID-19, it is essential to use pulse oximetry at the home setting to identify these patients.

## Introduction

Coronavirus disease (COVID-19) has been a mystery to the scientific world right from the discovery of the first case of COVID-19 pneumonia to its spread, presentation, and treatment. A baffling aspect of its presentation is hypoxia which might even be otherwise incompatible with life but without dyspnea, which is expected to occur to compensate for such a degree of hypoxia. This phenomenon is called silent hypoxia or happy hypoxia^1^. Since hypoxia in COVID-19 is an independent factor in predicting increased risk of intensive care unit requirement and in-hospital mortality, the presence of silent hypoxia as a presenting symptom in patients can be very treacherous as it might delay the diagnosis and subsequent initiation of treatment, giving the patient a false sense of well-being^2^. In the study of a cohort of 2080 patients at our center, the presence of hypoxia (SpO_2_<94%) was associated with 12 times higher odds of death^3^.

The incidence of silent hypoxia in COVID-19 has been reported from 32-65% in various studies^4–6^. The reports on patients with silent hypoxia is conflicting, with few studies reporting poorer outcomes while others report better outcomes^7^. In the pandemic setting, patients with SARS-CoV-2 infection are advised to isolate at home due to non-availability of hospital beds and seek hospitalization when red flags such as breathlessness and tachypnea occur.^8^ In patients with community acquired pneumonia, risk prediction tools such as CURB-65 and pneumonia severity index are used to decide if the patients need admission or can be managed on an outpatient basis. These scores rely on tachypnea to assess the respiratory function and do not recommend home pulse oximetry. This entity ‘silent hypoxia’ could be devastating if not recognized early by patients and caregivers, since they can be completely asymptomatic or present with only fever and upper respiratory symptoms without significant respiratory distress but may show severe hypoxia on pulse oximetry or blood gas analysis.

Physiologically, hypoxia causes stimulation of peripheral chemoreceptors present in the carotid body which signals the medulla oblongata to increase the minute ventilation and hence causesdyspnea^9^.Various theories have been put forward in attempts to explain the cause of silent hypoxia based on this phenomenon. Angiotensin-Converting Enzyme-2 receptors, which act as a receptor for entry of severe acute respiratory syndrome coronavirus 2 (SARS-CoV-2) virus into host cells, are also present in the carotid body. Hence, these receptors are implicated in causing a decrease in sensitivity of the carotid body to hypoxia leading to normal ventilation even in face of life-threatening hypoxia^10^. Also, SARS-CoV-2 infection leads to cytokine storm and neovascular proliferation in lungs which causes right to left shunting of blood and subsequently hypoxia. Hypoxia causes an increase in ventilation which leads to hypocapnia, as carbon dioxide is more diffusible than oxygen. The resulting hypocapnia will prevent any further increase in minute ventilation causing hypocapnic hypoxia without dyspnea^11^. Some theories suggest the spread of the virus from the oral cavity via a neural route through facial, glossopharyngeal, and vagus nerve to Nucleus Tractus Solitarius or from nasal cavity via the cribriform plate and ethmoidal sinus directly into the brain. Such spread might lead to inflammation and impaired signal processing of hypoxia at the higher centers leading to normal breathing despite severe hypoxia^12,13^. Other theories ranging from gut dysbiosis, formation of free radicals to impaired autonomic regulation have also been proposed^14–16^.

Though the physiological mechanism of of silent hypoxia is not very clear, it can potentially escalate to severe acute respiratory distress syndrome (ARDS), cardiorespiratory collapse, and even death as described in previous studies. We compare the clinical, laboratory and treatment parameters and evaluate the outcomes of patients with dyspneic and silent hypoxia in COVID-19 in a cohort of patients admitted to our hospital.

## Methods

### Study design

This was a retrospective cohort study conducted in the National Cancer Institute (Jhajjar), All India Institute of Medical Sciences, New Delhi, which is a tertiary care institute in India. The study was approved by the Institutional Ethics Committee of the Institution.

### Patients

We enrolled all consecutive patients who were admitted with SARS-CoV-2 infection at our institute who had hypoxia (SpO_2_< 94%) while breathing room air or needed oxygen support to maintain saturation >94%) at the time of presentation to the hospital. The demographic, clinical, and laboratory parameters of patients were collected from the case records and the electronic hospital management system of the hospital. All the patients included in our study were diagnosed SARS-CoV-2 infection by detecting viral RNA in respiratory samples by Reverse Transcription Polymerase Chain Reaction, Nucleic acid amplification tests or rapid antigen test.

### Case Definitions

Dyspnea is a subjective experience of breathing discomfort which was asked from the patient at the time of admission. Hypoxia was defined as oxygen saturation (SpO_2_) <94% on room air and severe hypoxia as SpO_2_<90% on room air^17,18^. Patients who were on oxygen supplementation to maintain a saturation above 94% were considered hypoxic regardless of oxygen saturation. The case definitions used in this study were based on the criteria described in the paper on the clinical features and outcomes of the entire cohort of patients who were treated at our institute during the period from April 2021 to June 2021^3^.

### Statistical analysis

The data were summarized using means and standard deviations for normal data and medians and interquartile ranges (p25-p75) for non-parametric data and means were compared using the ‘t test’ and medians using the Wilcoxon rank-sum test. The categorical data were summarized as proportions and compared using the Chi2 test or Fisher’s exact as appropriate. All statistical tests were performed with the use of a two-sided type I error rate of 5%. Missing data were not imputed and the summary parameters were calculated with the available data and the denominators (n) for each parameter was mentioned.

Univariate analysis was done to compare the various parameters between those who were discharged and those who died. Multivariate logistic regression analysis was done with models developed by including those that were found to be significant on univariate analysis as well as parameters of clinical relevance. We also included those parameters which we thought would influence the outcomes based on available scientific literature. Sensitivity analysis was done by dropping such parameters and by comparing the various models obtained by dropping them. All analysis was performed using STATA-Version 13.0 software.

## Results

Among 2080 COVID-19 patients admitted to our hospital from April to June 2021, 811 patients were presented with hypoxia. Among them, 637 patients (78.55%) had dyspnea (Dyspneic hypoxia group), and 174 patients (21.45%) had no dyspnea (Silent hypoxia group).The demographic and baseline characteristics among patients with dyspneic and silent hypoxia are compared in Table 1.

**Table 1:**
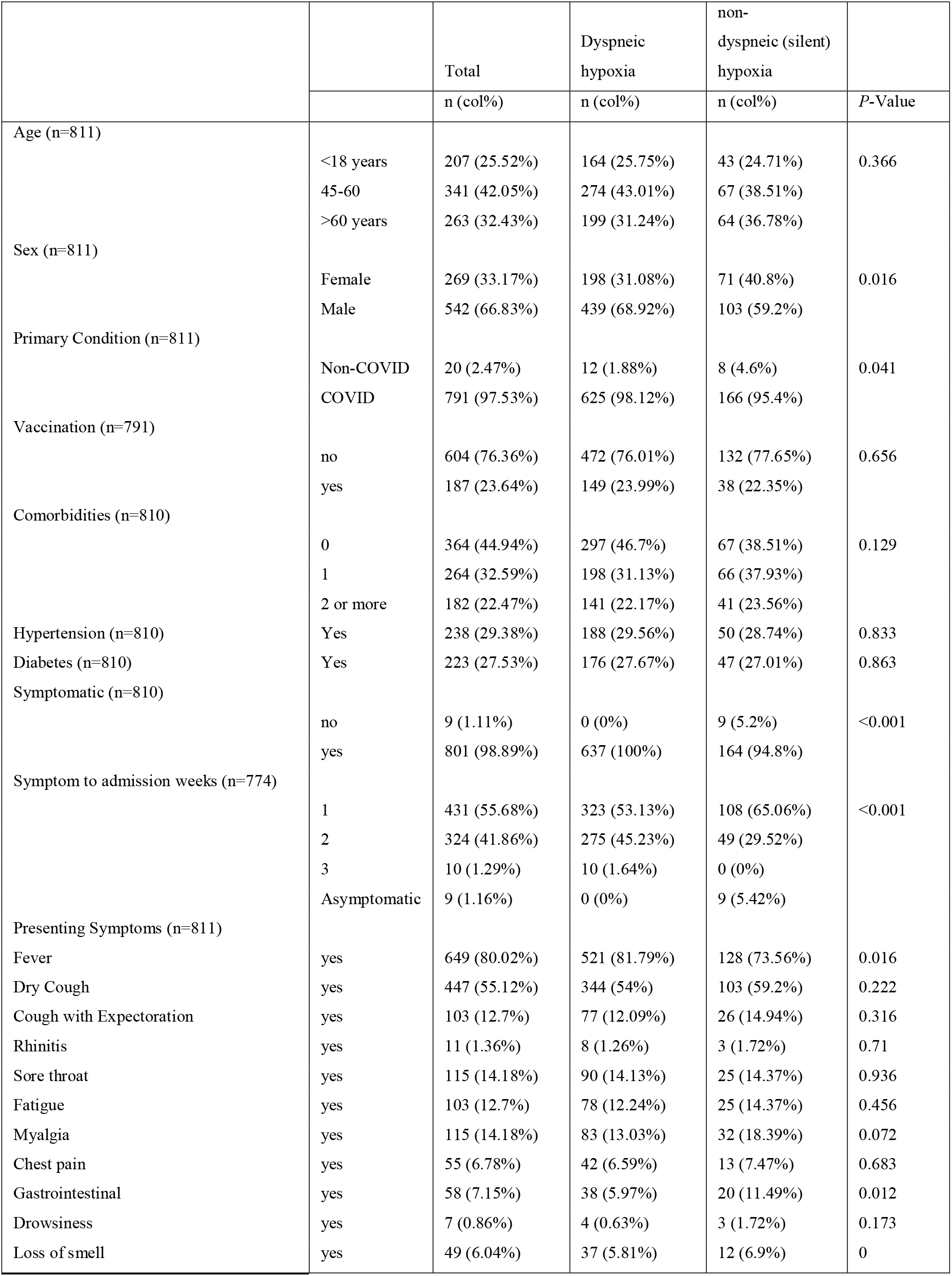
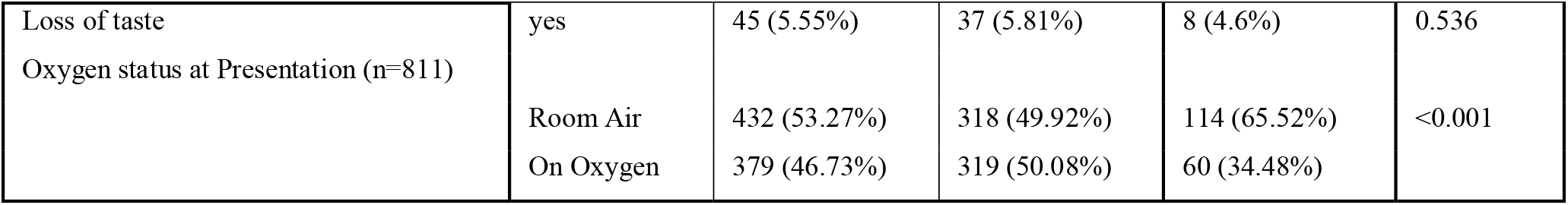
Demographic and baseline characteristics among patients with dyspneic and silent hypoxia

Out of the patients with silent hypoxia, 41% were males and 59% were females and this was statistically significant. It was found that 9 completely asymptomatic patients were hypoxic at the time of presentation to the screening area. This translates to 5.2% of cases presenting who were completely asymptomatic and hadhypoxia found only on pulse oximetry. Among the patients with silent hypoxia, 65% presented in the first week of symptoms when viremia plays a role in the pathogenesis as compared to 30% who presented in the second week during the inflammatory phase. This was in contrast to the patients with dyspneic hypoxia, in whom presentation in the inflammatory phase of the illness was higher (275 (45.2%) (p<0.001).

It is important to note that, almost half of the patients with dyspnea as a symptom along with hypoxia were brought to the hospital on oxygen. However, only 35% (60) of patients without dyspnea had their hypoxia diagnosed before reaching the hospital and been started on oxygen beforethe presentation by the paramedical workers during transportation (p<0.001). The rest of the demographic and clinical parameters were comparable between the patients with silent as well as dyspneic hypoxia. The laboratory parameters of these two groups are compared in Table 2. More patients with dyspneic hypoxia had leukocytosis (48.9%) as compared to silent hypoxia (33.6%) p=0.003. The rest of the laboratory parameters were comparable between the two groups. Table 3 compares the various treatments received by both groups. The high-frequency nasal cannula was used for oxygen delivery in 16% of patients with dyspneic hypoxia, while 9.5% were in the silent hypoxia group. High-dose methylprednisolone therapy was also given to a higher proportion of patients with dyspneic hypoxia as compared to silent hypoxia. Apart from these, no significant differences were seen between other treatment modalities such as antiviral drugs or tocilizumab between the groups. Multivariable logistic regression models (Table 4) were fitted to calculate the odds of death with silent hypoxia as the explanatory variable and other clinical, laboratory, and treatment parameters as covariates. We found that though these models showed a higher odds of death among patients with silent hypoxia, none of them were statistically significant.

**Table 2:** Laboratory parameters of patients with dyspneic and silent hypoxia

|  | Dyspneic Hypoxia |  | Non-Dyspneic (Silent) Hypoxia |  |  |
| --- | --- | --- | --- | --- | --- |
| Parameter | Available observations (N) | n(col%) | Available observations (N) | n(col%) | P-value |
| Anemia (Hb<11) | 520 | 87 (16.73%) | 140 | 26 (18.57%) | 0.608 |
| Leukocyte Count | 520 |  | 140 |  |  |
| • Normal |  | 252 (48.46%) |  | 85 (60.71%) | 0.003 |
| • Leukopenia (<4000/mm<sup>3</sup>) |  | 14 (2.69%) |  | 8 (5.71%) |  |
| • Leukocytosis (>11000/mm<sup>3</sup>) |  | 254 (48.85%) |  | 47 (33.57%) |  |
| Thrombocytopenia (<1.5lac/mm<sup>3</sup>) | 522 | 66 (12.64%) | 140 | 21 (15%) | 0.464 |
| Hyperferritinemia (>322 ng/mL) | 192 | 159 (82.81%) | 40 | 30 (75%) | 0.247 |
| D-dimer >500 ng/mL | 432 | 178 (41.2%) | 115 | 47 (40.87%) | 0.948 |
| IL-6 >4.4pg/mL | 328 | 267 (81.4%) | 68 | 58 (85.29%) | 0.446 |
| CRP >0.5 mg/dL | 485 | 474 (97.73%) | 133 | 128 (96.24%) | 0.337 |
| Creatinine>1 mg/dL | 528 | 124 (23.48%) | 146 | 44 (30.14%) | 0.1 |

\*WBC- White blood cell, LDH- Lactate dehydrogenase, SGPT- Serum glutamic pyruvic transaminase, ALT-Alanine aminotransferase, SGOT-Serum glutamic oxaloacetic transaminase, AST- Aspartate Aminotransferase, A:G – Albumin: Globulin, Hb- Hemoglobin, IL-6 – Interleukin-6

**Table 3:**
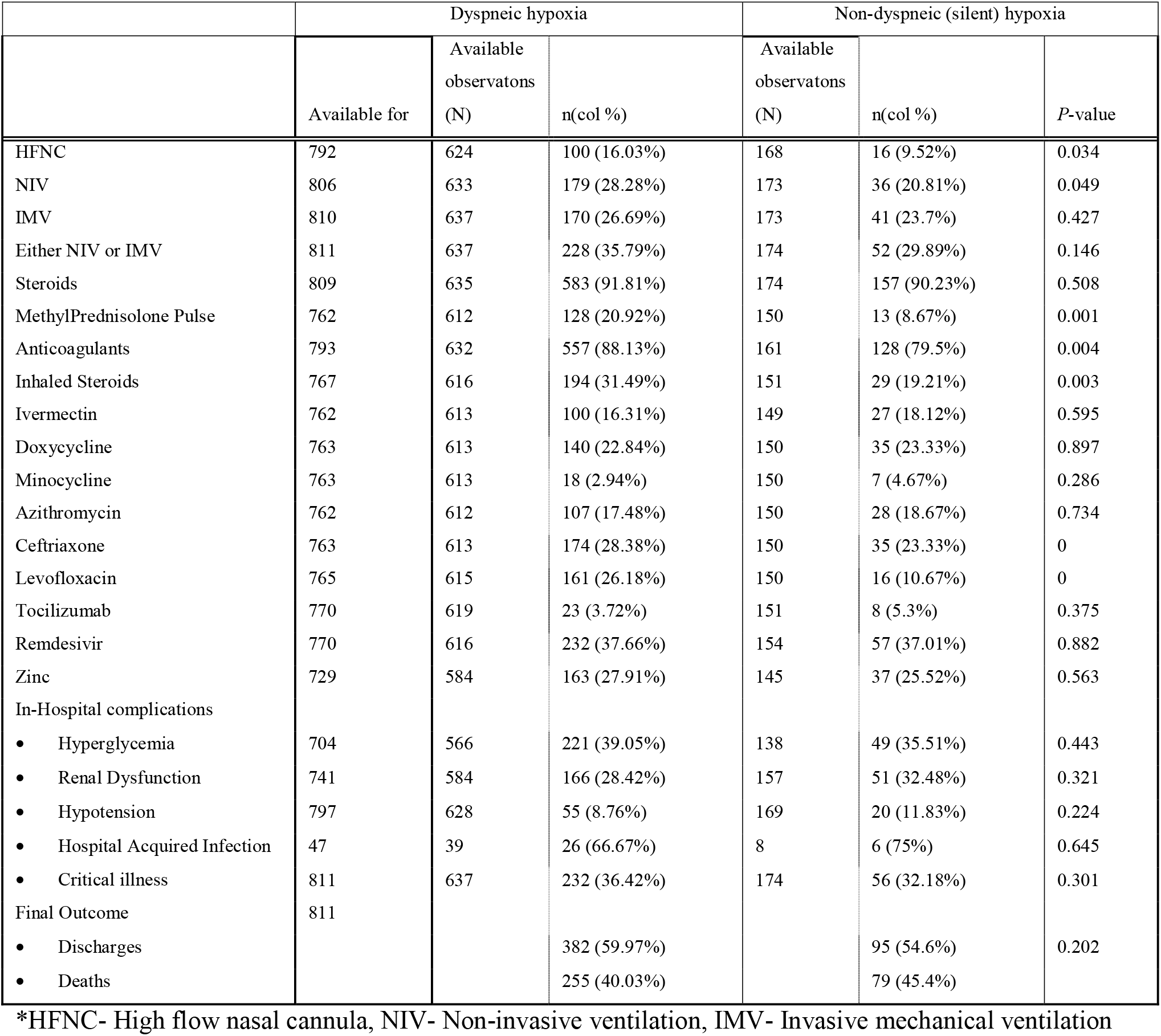
Treatment parameters of patients with dyspneic and silent hypoxia

**Table 4:** Odds of death among those with happy hypoxia compared to those without happy hypoxia

|  | Odds Ratio (95% CI) | *P*-Value |
| --- | --- | --- |
| Model 1: Univariate | 1.25 (0.89-1.75) | 0.202 |
| Model 2: Adjusted for age, gender, comorbidities, vaccination | 1.14 (0.8-1.64) | 0.474 |
| Model 3: Model 2 plus symptoms | 1.24 (0.81-1.89) | 0.323 |
| Model 4: Model 3 plus baseline lab parameters | 1.22 (0.68-2.19) | 0.504 |
| Model 5: Model 4 plus treatment parameters | 1.06 (0.43-2.61) | 0.894 |
| Model 6: Model 5 plus in-hospital complications | 1.1 (0.41-2.97) | 0.851 |

## Discussion

Though there are few case reports describing the perplexing entity – ‘silent hypoxia’, there are a handful of cohort studies which have described the demographic, clinical and laboratory findings in such patients. ^10,19,20^. Brouqui P et al^4^ in their retrospective study, analyzed data from March 3rd to April 27^th^, 2020 using dyspnea status, oxygen saturation, blood gas analysis, and low dose computed tomography scan reports. They defined hypoxia as SpO2≤95% They reported the incidence of silent hypoxia to be 14.2% based on oxygen saturation with pulse oximetry and 26.1% based on blood gas analysis. They reported these patients to be strongly associated with poor outcomes, the suggested cause being most patients belonging to the elderly age group and chronic diseases.

Another retrospective cohort study by Okuhama et al^5^, reported the incidence of silent hypoxia to be 3% and the authors defined hypoxia to be SpO2<94%. They reported that the patients with silent hypoxia might also have a poor prognosis but that however was not associated with old age or chronic diseases and suggested that some other mechanisms might be involved in this respect. This cohort did not have a comparison arm and was a descriptive study of 8 patients who presented with silent hypoxia amongst a total of 270 patients with COVID-19. None of these patients died, however, the authors did not compare these patients with silent hypoxia with those with dyspneic hypoxia, nor did they present the incidence of silent hypoxia amongst patients with hypoxia.

Busana et al^7^ reported a cohort of 213 patients with hypoxia defined as PaO2/FiO2 <300 as assessed by a blood gas analysis. (Partial pressure of oxygen/Fraction of inspired oxygen<300). They classified the patients into a dyspneic hypoxia group and silent hypoxia group and found that the mortality in the dyspneic group (29.7%) to be higher than that in the silent hypoxia group(17.6%) though these figures did not attain statistical significance (p=0.083).

Grimshaw et al^6^ reported a cohort of 470 patients with hypoxia defined as SpO2<80% and found that 5% of them had no breathlessness. In this study, the authors observed that the patients with silent hypoxia presented earlier to the hospital due to new onset headaches and the mortality was higher in patients with dyspneic hypoxia (43.2%) as compared to those with silent hypoxia (30.4%). The overall mortality in this cohort of was also higher than the reported mortality in other studies probably due to the definition of hypoxia to be a much lower oxygen saturation of <80%.

We defined hypoxia to be SpO2<94% as per the BTS guidelines for oxygen use^18^, Our study was done during the period from April to June 2021 during the ‘second wave’ of the pandemic in India. We found the incidence of silent hypoxia to be 21.45%. Our study did not find any statistically significant difference in outcome between the silent and dyspneic hypoxic groups. Also, the age and comorbid disease status were not significantly different in our patients suggesting that silent hypoxia was a presentation that was equally spread among all ages and co-morbidities in the population. ^21^

We found leukocytosis to be significantly more in patients with dyspneic hypoxia. Since leukocytosis points towards a hyperinflammatory state, further research is required to know if dyspneic hypoxia has different pathophysiology as compared to patients with silent hypoxia^22^. The presence of leukocytosis could have led to these patients being identified as having a cytokine storm and thus could have led the clinicians to significantly greater use of Methylprednisolone pulse therapy, inhaled corticosteroids, antibiotics, and also respiratory support like Non-invasive ventilation (NIV) and High-flow nasal cannula (HFNC) in the dyspneic hypoxic group. A more aggressive approach in treatment was adopted in the dyspneic hypoxic group in terms of using high dose methylprednisolone pulse but the usage of anti interleukin therapy such as tocilizumab or antivirals were comparable between the groups. In patients with baseline hypoxia, in both groups, which later deteriorated and progressed to death, inflammatory markers such as d-dimer wereelevated in more than 40% of patients while IL-6 levels were elevated in more than 80% of patients. This re-enforces the fact that it has a strong correlation with disease severity and is a reliable prognostic marker for in-hospital mortality in patients with COVID-19^23^. However, these inflammatory markers did not differ between the patients with silent and dyspneic hypoxia in our cohort.

In a comparative study of vaccinated and unvaccinated individuals from our cohort, we found that vaccination significantly reduced the odds of developing hypoxia and death^24^. However the presentation of silent hypoxia was not significantly different in the groups receiving the vaccination against COVID-19 and nonvaccinated groups. Further research on the pathogenesis of post-vaccination breakthrough infections is needed.In our cohort, 21.45% of patients with hypoxia did not have breathlessness and were found to be hypoxic only on pulse oximetry. Since it is well known that the presence of hypoxia is the strongest predictor of death, this finding emphasizes the fact that in addition to clinical examination, pulse oximetry should be an integral part of disease assessment at the primary care level and mere presence or absence of dyspnea should not be used to triage patients. Since pulse oximetry is an easy to use non-invasive method to watch for silent hypoxia at home, the COVID-19 patients undergoing home isolation should be suggested to undergo such monitoring regularly, for early diagnosis and seeking treatment before its too late^25^. The general public should also be educated that silent hypoxia is also a presenting feature of COVID 19 and they should look for an increase in respiratory rate without any discomfort to the patient so that presence of such features does not go unrecognized. As observed in this cohort, more patients who had dyspneic hypoxia were started on oxygen by the paramedical personnel as compared to those with silent hypoxia. This data suggests that the presence of dyspnea, is easily triaged, and silent hypoxia might be missed by the public as well as paramedics. Since silent hypoxia is similar to dyspneic hypoxia in terms of outcomes, it is imperative to do pulse oximetry in every patient diagnosed with SARS-CoV-2 infection. The major limitation of the present study is its retrospective nature. Some parameters were missing in the collected data. We have excluded those parameters from our study analysis. We retrospectively obtained pulse oximetry and breathlessness data that is subject to bias. Our findings, however, may be useful for understanding more about silent hypoxia.

## Conclusion

Silent hypoxia is a significant problem in COVID-19. Hypoxia if untreated can be fatal and with huge numbers of patients during each ‘wave’, patients with dyspneic hypoxia are more likely to visit the hospital and get triaged into severe illness categories and thus receive quick and appropriate care. On the other hand, patients with silent hypoxia, are likely to just get tested due to other symptoms only to be found to be hypoxic later, or be completely unaware of the disease and found only on pulse oximetry or blood gas analysis. However, once the patient lands in the hospital, both dyspneic and silent hypoxia, have a similar clinical course, and silent hypoxia does not seem to alter the natural history among hospitalized patients with COVID-19. Since silent hypoxia and dyspneic hypoxia have similar outcomes, it is of paramount importance to do pulse oximetry in every patient diagnosed with SARS-CoV-2 infection.

## Data Availability

Data will be made available to others upon reasonable request to be routed
through our Institute ethics committee with an appropriate protocol

## Abbreviations

COVID-19: Coronavirus disease
HFNC: High flow nasal cannula
IMV: Invasive mechanical ventilation
NIV: Non-invasive ventilation
SARS-CoV-2: severe acute respiratory syndrome coronavirus 2
SpO_2_: Peripheral oxygen saturation

## References

1. Bickler PE, Feiner JR, Lipnick MS, McKleroy W. “Silent” Presentation of Hypoxemia and Cardiorespiratory Compensation in COVID-19. Anesthesiology 2021;134(2):262–269.

2. Xie J, Covassin N, Fan Z, et al. Association Between Hypoxemia and Mortality in Patients With COVID-19. Mayo Clin Proc 2020;95(6):1138–1147.

3. Elavarasi A, Sagiraju HKR, Garg RK, et al. Clinical features, demography and predictors of outcomes of SARS-CoV-2 infection in a tertiary care hospital in India-A cohort study [Internet]. Infectious Diseases (except HIV/AIDS); 2021 [cited 2021 Aug 24]. Available from: http://medrxiv.org/lookup/doi/10.1101/2021.08.10.21261855

4. Brouqui P, Amrane S, Million M, et al. Asymptomatic hypoxia in COVID-19 is associated with poor outcome. Int J Infect Dis 2021;102:233–238.

5. Okuhama A, Ishikane M, Hotta M, et al. Clinical and radiological findings of silent hypoxia among COVID-19 patients. Journal of Infection and Chemotherapy 2021;27(10):1536–1538.

6. García-Grimshaw M, Flores-Silva FD, Chiquete E, et al. Characteristics and predictors for silent hypoxemia in a cohort of hospitalized COVID-19 patients. Autonomic Neuroscience 2021;235:102855.

7. Busana M, Gasperetti A, Giosa L, et al. Prevalence and outcome of silent hypoxemia in COVID-19. Minerva Anestesiol [Internet] 2021 [cited 2021 Aug 26];87(3). Available from: https://www.minervamedica.it/index2.php?show=R02Y2021N03A0325

8. World Health Organization. Clinical management of severe acute respiratory infection (SARI) when COVID-19 disease is suspected. Interim guidance. Pediatr Med Rodz 2020;16(1):9–26.

9. Moosavi SH, Golestanian E, Binks AP, Lansing RW, Brown R, Banzett RB. Hypoxic and hypercapnic drives to breathe generate equivalent levels of air hunger in humans. J Appl Physiol (1985) 2003;94(1):141–154.

10. Tobin MJ, Laghi F, Jubran A. Why COVID-19 Silent Hypoxemia Is Baffling to Physicians. Am J Respir Crit Care Med 2020;202(3):356–360.

11. Jounieaux V, Rodenstein DO, Mahjoub Y. On Happy Hypoxia and on Sadly Ignored “Acute Vascular Distress Syndrome” in Patients with COVID-19. Am J Respir Crit Care Med 2020;202(11):1598–1599.

12. U R A, Verma K. Happy Hypoxemia in COVID-19-A Neural Hypothesis. ACS Chem Neurosci 2020;11(13):1865–1867.

13. Nouri-Vaskeh M, Sharifi A, Khalili N, Zand R, Sharifi A. Dyspneic and non-dyspneic (silent) hypoxemia in COVID-19: Possible neurological mechanism. Clin Neurol Neurosurg 2020;198:106217.

14. Gopal AB, Chakraborty S, Padhan PK, et al. Silent hypoxia in COVID-19: a gut microbiota connection. Curr Opin Physiol 2021;23:100456.

15. Baker J, Incognito AV, Wilson RJA, Raj SR. Syncope and silent hypoxemia in COVID-19: Implications for the autonomic field. Auton Neurosci 2021;235:102842.

16. Machado-Curbelo C. Silent or “Happy” Hypoxemia: An Urgent Dilemma for COVID-19 Patient Care. MEDICC Rev 2020;22(4):85–86.

17. CLINICAL GUIDANCE FOR MANAGEMENT OF ADULT COVID-19 PATIENTS | AIIMS Covid Information Portal [Internet]. [cited 2021 Aug 26];Available from: https://covid.aiims.edu/clinical-guidance-for-management-of-adult-covid-19-patients/

18. O’Driscoll BR, Howard LS, Earis J, Mak V. BTS guideline for oxygen use in adults in healthcare and emergency settings. Thorax 2017;72(Suppl 1):ii1–ii90.

19. Lari A, Alherz M, Nouri A, Botras L, Taqi S. Caution against precaution: A case report on silent hypoxia in COVID-19. Ann Med Surg (Lond) 2020;60:301–303.

20. Chandra A, Chakraborty U, Pal J, Karmakar P. Silent hypoxia: a frequently overlooked clinical entity in patients with COVID-19. BMJ Case Rep 2020;13(9):e237207.

21. Mocanu A, Noja GG, Istodor AV, et al. Individual Characteristics as Prognostic Factors of the Evolution of Hospitalized COVID-19 Romanian Patients: A Comparative Observational Study between the First and Second Waves Based on Gaussian Graphical Models and Structural Equation Modeling. J Clin Med 2021;10(9):1958.

22. Wang J, Jiang M, Chen X, Montaner LJ. Cytokine storm and leukocyte changes in mild versus severe SARS-CoV-2 infection: Review of 3939 COVID-19 patients in China and emerging pathogenesis and therapy concepts. J Leukoc Biol 2020;108(1):17–41.

23. Yao Y, Cao J, Wang Q, et al. D-dimer as a biomarker for disease severity and mortality in COVID-19 patients: a case control study. j intensive care 2020;8(1):49.

24. Sagiraju HKR, Elavarasi A, Gupta N, et al. The effectiveness of SARS-CoV-2 vaccination in preventing severe illness and death – real world data from a cohort of patients hospitalized with COVID-19. MedRxiv 2021

25. Jouffroy R, Jost D, Prunet B. Prehospital pulse oximetry: a red flag for early detection of silent hypoxemia in COVID-19 patients. Crit Care 2020;24(1):313.

